# Persistent or recurrent depression in young adults: associations with systemic inflammation and early life adversity

**DOI:** 10.64898/2026.11.10.26367713

**Authors:** Freya R. Shepherd, Sophie J. Fairweather, Laura D. Howe, Hannah J. Jones, Golam M. Khandaker

## Abstract

1

**Background:** There is limited research on persistent or recurrent depression into young adulthood. We examined the prevalence of recurrent/persistent depression in young adults, and associations with childhood trauma/adversity and systemic inflammation, and if inflammatory markers mediated associations between childhood trauma and subsequent depression.

**Methods:** ALSPAC participants with ICD-10 depression assessments at ages 18, 24 and 30 were classified as never depressed, transiently depressed (one episode), or persistently/recurrently depressed (≥2 episodes). Trauma/adversity from birth to age 16/17 was characterised by type, timing and burden. Interleukin-6 (IL-6) and C-reactive protein (CRP) were measured at ages 9, 16, 18 and 24. Analyses used multinomial logistic regression, linear regression and causal mediation models.

**Results:** Among 2,014 participants, 78.9% were never depressed, 15.0% had transient depression and 6.1% had persistent/recurrent depression. Trauma/adversity was more strongly associated with persistent/recurrent depression (RRR 3.17, 95% CI 1.98-5.06; FDR-P=6.2×10⁻⁶) than transient depression, with a dose-response pattern and stronger associations for threat adversities. Middle-to-late childhood (11-17 yrs) emerged as a sensitive period for persistent/recurrent (RRR 2.85, 95% CI 1.47-5.52) and transient depression (RRR 1.85, 95% CI 1.25-2.73). Trauma/adversity was associated with modestly higher IL-6 at age 24 (all FDR-adjusted P<0.05); CRP associations did not survive multiple-testing correction. Neither marker mediated trauma/adversity associations with persistent/recurrent depression.

**Conclusion:** Persistent/recurrent depression affects approximately 1 in 20 young adults. Childhood trauma/adversity, particularly during middle-to-late childhood, was associated with persistent/recurrent depression, but measured inflammatory proteins did not mediate this association. Further work is needed to understand biological mechanisms underlying the embedding early life psychosocial stress.

## 2 Introduction

Depression commonly emerges in early life and often follows a chronic, relapsing-remitting course. Approximately 35% of mood disorders begin before age 25 (Solmi et al., 2022), with depression typically emerging earlier (Bromet et al., 2011). Persistent, recurrent depression during adolescence to early adulthood imposes significant personal and clinical burden, including impaired social, educational, and occupational functioning (Thapar et al., 2022). However, the prevalence of persistent/recurrent depression in young adults during the second and third decades of life remain poorly characterised. It is also unclear how established psychosocial and biological risk factors, such as childhood trauma/adversity and inflammation (Mac Giollabhui et al., 2025; Nelson et al., 2017; Watson et al., 2025), for depression relate to persistent/recurrent depression into adulthood.

Childhood trauma/adversity, often assessed using adverse childhood experience (ACE) measures (Felitti et al., 1998), has a strong, dose-response relationship with the risk and severity of depression throughout life (Flouri et al., 2020; Iob et al., 2022; LeMoult et al., 2020; Nanni et al., 2012; Tan & Mao, 2023). Beyond cumulative exposure, the nature and developmental timing of adversity may influence subsequent depression. Threat-related adversities have shown stronger associations with elevated depressive symptom trajectories than other ACEs (Iob et al., 2022) and maltreatment has been associated with a more persistent/recurrent course of depression with poorer treatment outcomes (Nanni et al., 2012). However, the extent to trauma/adversity type and timing are differentially associated with transient or persistent/recurrent depression into adulthood remains unclear.

Altered systemic inflammation may link ACEs with the development and persistence of depression. A meta-analysis of longitudinal studies associated higher circulating interleukin-6 (IL-6) and C-reactive protein (CRP) concentrations with subsequent depressive symptoms, although associations were small (Mac Giollabhui et al., 2021). In the Avon Longitudinal Study of Parents and Children (ALSPAC), higher childhood IL-6 was associated with depression at age 18 (Khandaker et al., 2014), while higher IL-6 at age 9 was associated with persistent depressive symptoms between ages 10 and 19, but not adolescent-onset symptoms (Khandaker et al., 2018). These findings suggest that inflammation may precede depression and be particularly relevant to persistent symptoms, supporting investigation of its potential mediating role between ACEs and depression.

Evidence linking childhood trauma/adversity to systemic inflammation is mixed, with associations varying by adversity type, sex, inflammatory marker and age at assessment. Lacey et al. reported that parental separation or divorce was associated with higher IL-6, whereas several other adversities were associated with lower IL-6, with further differences according to the sex of the child (Lacey et al., 2020). Crick et al. found no evidence of associations between cumulative or individual ACEs and inflammatory marker glycoprotein acetyls (GlycA) from ages 8 to 24, but observed a small association between cumulative ACE exposure and GlycA among mothers at age 49 (Crick et al., 2022). Similarly, older adults reporting three or more ACEs retrospectively had chronically elevated CRP compared with those reporting none (Iob et al., 2020). These findings suggest that inflammatory correlates of adversity may differ across the life course, although their biological significance remains uncertain. Further investigation is therefore needed to establish whether adversity-related inflammation helps explain transient or persistent/recurrent depression across young adulthood.

Despite evidence linking childhood adversity to depression, and separate evidence implicating inflammation in depression risk and further separate evidence linking trauma/adversity to inflammatory disturbances, relatively little is known about how trauma exposure, inflammatory biomarkers, and depression course are related within the same longitudinal framework into early adulthood. Inflammation may mediate the relationship between trauma/adversity and later depression, but current evidence is limited and focuses on IL-6 in childhood (up to age 10) (Flouri et al., 2020). Repeated assessments extending into the third decade of life provide an opportunity to distinguish depression identified at a single assessment from depression present at multiple assessments, and to examine whether these patterns share early-life risk factors. Understanding these associations could inform efforts to prevent enduring depression and clarify potential biological pathways linking childhood adversity with later mental health.

Using data from ALSPAC, a population-based prospective birth cohort, we have examined the longitudinal course of depression in young adults based on ICD-10 diagnosis of depression at ages 18, 24 and 30 years identifying those with transient (one episode) and persistent/recurrent (two or more episodes) depression. We have examined the associations of transient and persistent/recurrent depression with: i) childhood trauma/adversity including the impact of specific trauma types, cumulative exposure, and developmental timing of trauma exposure; and ii) systemic inflammatory marker levels in blood from childhood through to adulthood including any mediating effects of inflammation on the trauma-depression relationship.

## 3 Methods

### 3.1 Data Source

This study uses data from the ALSPAC cohort. Pregnant women resident in Avon, UK with expected dates of delivery between 1st April 1991 and 31^st^ December 1992 were invited to take part in ALSPAC. The initial number of pregnancies enrolled was 14,541. Of these initial pregnancies, 13,988 children were alive at 1 year of age. When the children were approximately 7 years old, an attempt was made to expand the initial sample by recruiting eligible participants who had not originally enrolled. The total sample available for analyses using data collected after age 7 increased to 15,447 pregnancies, resulting in 15,658 foetuses. Of these, 14,901 children were alive at 1 year of age (Boyd et al., 2013; Fraser et al., 2013; Northstone et al., 2019, 2026). Study data were collected and managed using REDCap electronic data capture tools hosted at the University of Bristol (Harris et al., 2009). REDCap (Research Electronic Data Capture) is a secure, web-based software platform designed to support data capture for research studies. Please note that the study website contains details of all the data that is available through a fully searchable data dictionary and variable search tool (http://www.bristol.ac.uk/alspac/researchers/our-data/). Ethical approval for the study was obtained from the ALSPAC Ethics and Law Committee and the Local Research Ethics Committees. Informed consent for the use of all data collected was obtained from participants following the recommendations of the ALSPAC Ethics and Law Committee at the time. Participants can contact the study team at any time to retrospectively withdraw consent for their data to be used. Study participation is voluntary and during all data collection sweeps, information was provided on the intended use of data.

### 3.2 Assessment of trauma and adverse childhood experiences

Childhood trauma/adversity was assessed using two sets of variables available in ALSPAC: trauma variables and adverse childhood experience (ACE) variables. Using both sets enabled us to include a broader range of specific adversities while also examining exposure during distinct developmental periods. Where exposures overlapped, trauma variables were used because they provided period-specific measures. All included variables are described in **Supplementary Table 1**.

Croft *et al*. derived trauma variables from 121 questions across parent-completed questionnaires and participant self-reports (Croft et al., 2019). These captured any trauma and specific trauma types from birth to age 17 and within early childhood (0-5 yrs), middle childhood (5-11 yrs), and adolescence (11-17 yrs). Types included physical, sexual, and emotional abuse; emotional neglect; domestic violence; and bullying.

Houtepen *et al*. derived ACE variables from 541 questionnaire items covering ages 0-16 years (Houtepen et al., 2018). These captured ten classic ACE categories: sexual, physical, and emotional abuse; emotional neglect; parental substance misuse; parental mental illness or suicide attempt; violence between parents; parental separation; bullying; and parental criminal conviction. Data before age eight were predominantly parent-reported; participants also provided self-reports thereafter.

### 3.3 Assessment of systemic inflammatory markers

Levels of CRP were measured in blood samples collected at ages 9, 16, 18 and 24 years, and IL-6 levels were measured in blood samples collected at ages 9 and 24 (see **Supplementary Methods 1** for further information). CRP concentrations (mg/L) were measured using ELISA and standardised to Z-scores prior to analysis. IL-6 (pg/mL) was also measured via ELISA and transformed at all age points, other than at age 24 (where no ELISA IL-6 variable is available) where levels were measured via Olink assay and expressed as NPX values on a log2 scale. At each assessment, CRP values >10 mg/L were set to missing for that timepoint only, before z-transformation, to reduce the influence of elevations potentially reflecting acute infection or other inflammatory conditions (Pearson et al., 2003).

### 3.4 Assessment of depression and definition of transient, and persistent/recurrent depression

Depression was assessed at ages 18, 24, and 30 years using the clinical interview schedule revised (CIS-R) (Lewis et al., 1992). CIS-R is a computerised, self-administered, semi-structured interview which assesses symptoms of common mental disorders occurring in the past two weeks and provides diagnosis of depression and other common mental disorders according to ICD-10 criteria. The CIS-R has been extensively validated and used for largescale epidemiological research and RCTs such as the NHS England Adult Psychiatric Morbidity Survey (Brugha et al., 1999; McManus et al., 2016).

Longitudinal depression groups were defined using ICD-10 depression diagnoses at ages 18, 24, and 30 years. Participants were classified as never depressed (no diagnosis at any assessment), transiently depressed (a diagnosis at one assessment only), or persistently/recurrently depressed (a diagnosis at two or more assessments).

The main analysis included participants with depression status available at all three assessments and those whose longitudinal classification could be determined despite missing data. Specifically, participants with depression at two assessments and missing status at the third were classified as persistently/recurrently depressed, as the missing value could not alter their classification (see **Supplementary Methods 2**).

ICD-10 depression diagnoses at ages 18, 24, and 30 years were also examined as separate binary outcomes, with participants classified as depressed or not depressed at each assessment.

### 3.5 Assessment of potential confounders

Potential confounders included sex, ethnicity, maternal age at delivery, maternal education and maternal social class. Biomarker analyses were additionally adjusted for blood collection variables (month of collection, time of collection and infection status). See **Supplementary Methods 3** for further information.

### 3.6 Statistical Analysis

Analyses were conducted in RStudio version 4.6.0. Analysis code is available at GitHub in public repository (https://github.com/shepherdfreya/immunometabolic_depression).

Missing covariate data were imputed under a missing at random assumption using multiple imputation by chained equations, generating 20 imputed datasets (list of auxiliary methods used in **Supplementary Table 1**). Auxiliary sociodemographic, biological, and timing-related variables were included as predictors but were not imputed. Further details of the imputation procedure are provided in **Supplementary Methods.**

Results are presented as odds ratios (ORs) or relative risk ratios (RRRs) with 95% confidence intervals and p-values. RRRs represent the relative change in risk associated with each one-point increase in the predictor score. For analyses of depression trajectory groups, the Never group was used as the reference category, with estimates presented for Transient versus Never and Persistent/Recurrent versus Never. Models were fitted within each imputed dataset and pooled using Rubin’s rules. P-values were adjusted for multiple testing using false discovery rate (FDR) correction within each analysis, model, and outcome contrast.

#### 3.6.1 Associations of adversity/trauma with depression and inflammatory markers

Regression analyses examined associations of childhood ACEs and trauma with depression and circulating inflammatory markers. Multinomial logistic regression modelled longitudinal depression groups (reference: never depressed), binary logistic regression modelled depression at individual ages, and linear regression modelled CRP at ages 9, 16, 18 and 24 and IL-6 at ages 9 and 24 years.

Exposures included any trauma, cumulative ACE and trauma scores, individual adversity types across childhood, and trauma within developmental periods (0-5, 5-11 and 11-17 yrs).

Models were unadjusted and adjusted for sex, child ethnicity, maternal age at delivery, maternal education and maternal social class. Inflammatory marker models additionally adjusted for sample month, sample time and infection before the visit.

Inflammatory markers were measured between ages 9 and 24, whereas depression trajectories covered ages 18, 24 and 30. To preserve temporal ordering, biomarker-depression analyses used the nearest subsequent binary depression assessment.

#### 3.6.2 Mediation analysis

We tested whether IL-6 or CRP at age 24 mediated associations between childhood adversity/trauma and ICD-10 depression at age 30. Exposure measures were selected based on associations with IL-6 that survived FDR correction; corresponding CRP mediation pathways were examined as exploratory analyses.

Causal mediation analyses were conducted using the parametric g-formula implemented in CMAverse. Models adjusted for child sex and ethnicity, maternal age at delivery, maternal education, maternal occupational social class, and blood collection month and time at age 24. BMI at age 24 was included as an intermediate confounder.

We estimated total effects, direct and indirect effects, and the corresponding proportion mediated. Models were fitted separately within each of 20 imputed datasets, with 1,000 bootstrap replications within each dataset.

#### 3.6.3 Sensitivity analysis

Sensitivity analyses repeated biomarker analyses including participants with CRP concentrations greater than 10 mg/L. Analyses were also repeated using complete observed covariate data only, excluding imputed covariate values. Finally, analyses of longitudinal depression groups were repeated among participants with depression status available at all three assessments, excluding those classified as persistently/recurrently depressed based on two observed diagnoses and one missing assessment. See **Supplementary Methods 5** for further description.

## 4 Results

### 4.1 Prevalence of persistent/recurrent depression diagnosis at ages 18, 24 and 30 years and associations with sociodemographic factors

Among all participants with valid ICD-10 depression status at each assessment, 355 of 4,513 (7.9%) met criteria for depression at age 18, compared with 340 of 3,852 (8.8%) at age 24 and 324 of 3,614 (9.0%) at age 30 (**Supplementary Table 3**). These age-specific samples included participants regardless of whether depression status was available at other assessments.

The longitudinal analysis included 2,014 participants: 1,966 with depression status available at all three assessments and 48 with depression at two assessments and missing status at the third, allowing unambiguous classification as persistently/recurrently depressed. Within this longitudinal sample, 1,590 (78.9%) were classified as never depressed, 302 (15.0%) as transiently depressed, and 122 (6.1%) as persistently/recurrently depressed (**Figure 1**).

**Figure 1.**
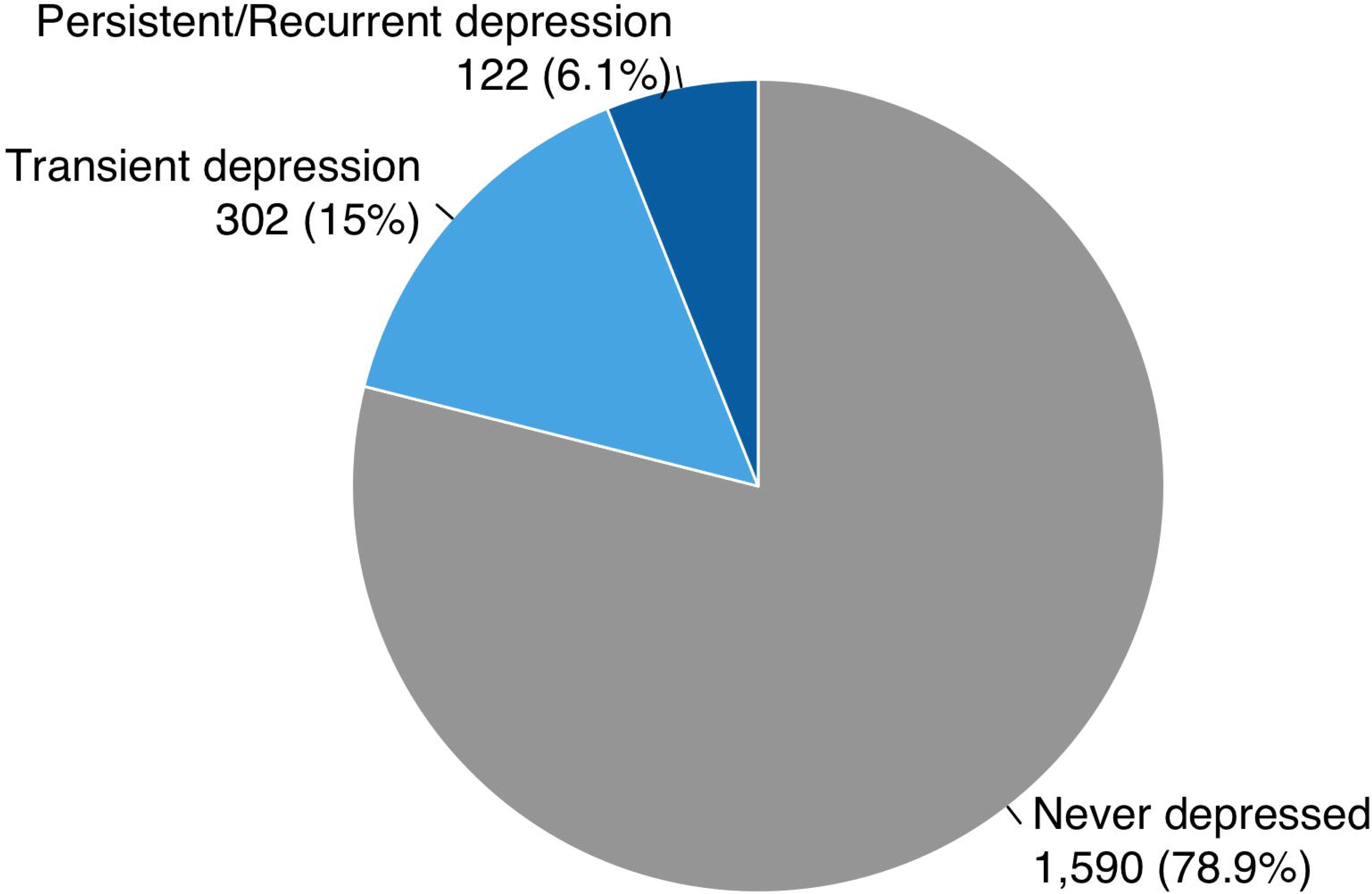
Longitudinal depression patterns across ages 18, 24, and 30 years in the ALSPAC birth cohort. Note: Among 2,014 participants in the longitudinal analysis, 1,590 (78.9%) were classified as never depressed, 302 (15.0%) as transiently depressed (depression at one assessment only), and 122 (6.1%) as persistently/recurrently depressed (depression at two or more assessments). The sample included 1,966 participants with depression status available at all three assessments and 48 with depression at two assessments and missing status at the third.

Several sociodemographic characteristics differed across depression groups, including sex, maternal education, marital status and housing type (**Supplementary Table 4**). Persistent/recurrent depression was associated with female sex and mothers with lower education achievement, divorced or never married, living in council rented accommodation.

### 4.2 Associations of persistent/recurrent depression with childhood adversity/trauma

Results are summarised in **Figure 2** and **Supplementary Table 7.** Overall, childhood trauma/adversity was consistently associated with increased risk of subsequent depression, with stronger associations observed for persistent/recurrent depression than transient depression.

**Figure 2.**
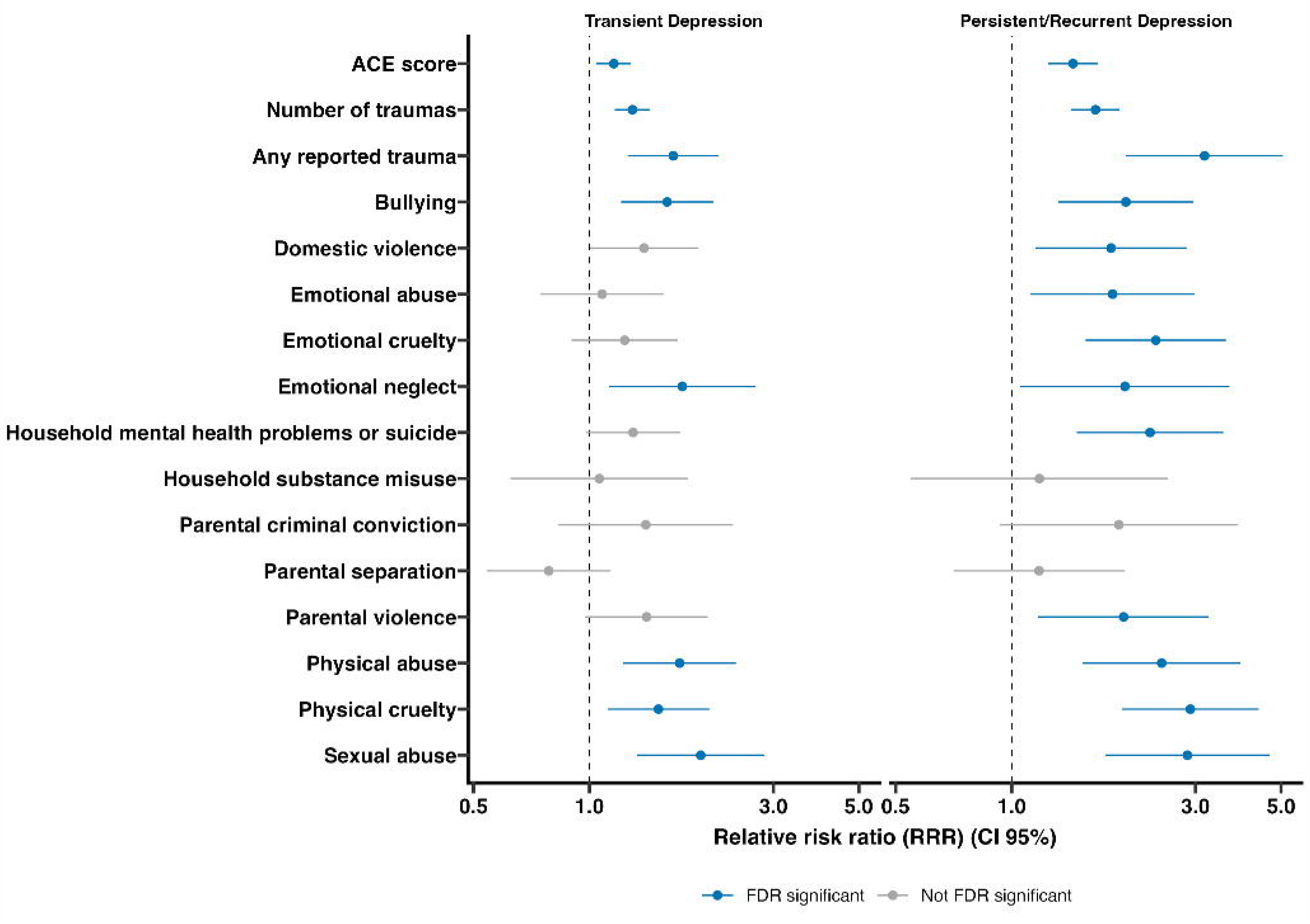
Associations of childhood adversity/trauma with transient depression (one episode), persistent or recurrent depression (two or more episodes) across ages 18, 24 and 30 years. Note: Relative risk ratios (RRRs) and 95% confidence intervals (CIs) represent increase in risk of transient or persistent/recurrent depression diagnosis across ages 18, 24, 30 years. The exposures are childhood adversity/trauma coded as continuous (number of traumas / ACE score) or binary (all other exposures) variable. Never depressed was used as the reference category. Regression models have been adjusted for sex, ethnicity, maternal age at delivery, maternal education, and maternal social class. Blue represents associations surviving false discovery rate (FDR) correction, while grey associations non surviving FDR correction.

Individuals exposed to any childhood trauma/adversity had over three times the risk of persistent/recurrent depression compared with those reporting no trauma (RRR 3.17, 95% CI 1.98-5.06, FDR-P = 6.2 × 10⁻⁶), while the association with transient depression was more modest (RRR 1.65, 95% CI 1.26-2.16, FDR-P = 2.46 × 10⁻^3^).

Among individual trauma/adversity types, threat related types had the strongest associations with depression. Physical cruelty (RRR 2.91, 95% CI 1.93-4.37, FDR-P = 2.25 × 10⁻⁶), sexual abuse (RRR 2.86, 95% CI 1.75-4.67, FDR-P = 9.07 × 10⁻^5^), physical abuse (RRR 2.45, 95% CI 1.53-3.92, FDR-P = 4.29 × 10⁻^4^) and emotional cruelty (RRR 2.37, 95% CI 1.55-3.59, FDR-P = 1.49 × 10⁻^5^) were all associated with a substantially increased risk of persistent/recurrent depression. These associations were consistently stronger than those observed for transient depression, although sexual abuse, physical abuse and physical cruelty also remained significantly associated with transient depression.

Continuous measures of cumulative trauma/adversity (ACE score and trauma count) showed evidence of dose-response in their associations with depression. Each additional reported trauma increased the risk of persistent/recurrent depression by 65% (RRR 1.65, 95% CI 1.43-1.91, FDR-P = 2.24 x 10⁻^10^), while each additional ACE score points increased risk by 44% (RRR 1.44, 95% CI 1.24-1.68, FDR-P = 6.2 x 10^-6^). Associations were also evident for transient depression, although effect sizes were consistently smaller. Adjustment for sex, ethnicity, maternal age, maternal education, and maternal social class had little impact on the observed associations, with adjusted and unadjusted models yielding very similar results.

### 4.3 Age of trauma exposure identifies middle and late childhood as sensitive periods for persistent/recurrent depression in adulthood

To investigate whether there is a sensitive period during which trauma is more strongly associated with subsequent depression, trauma exposure was examined across three developmental periods: 0-5, 5-11 and 11-17 yrs, using both binary and continuous measures (**Figure 3**, **Supplementary Table 8**). Overall, trauma exposure during middle childhood (5-11 yrs) and adolescence (11-17 yrs) was associated with later depression, whereas there was little evidence of an association for exposure during early childhood (0-5 yrs).

**Figure 3.**
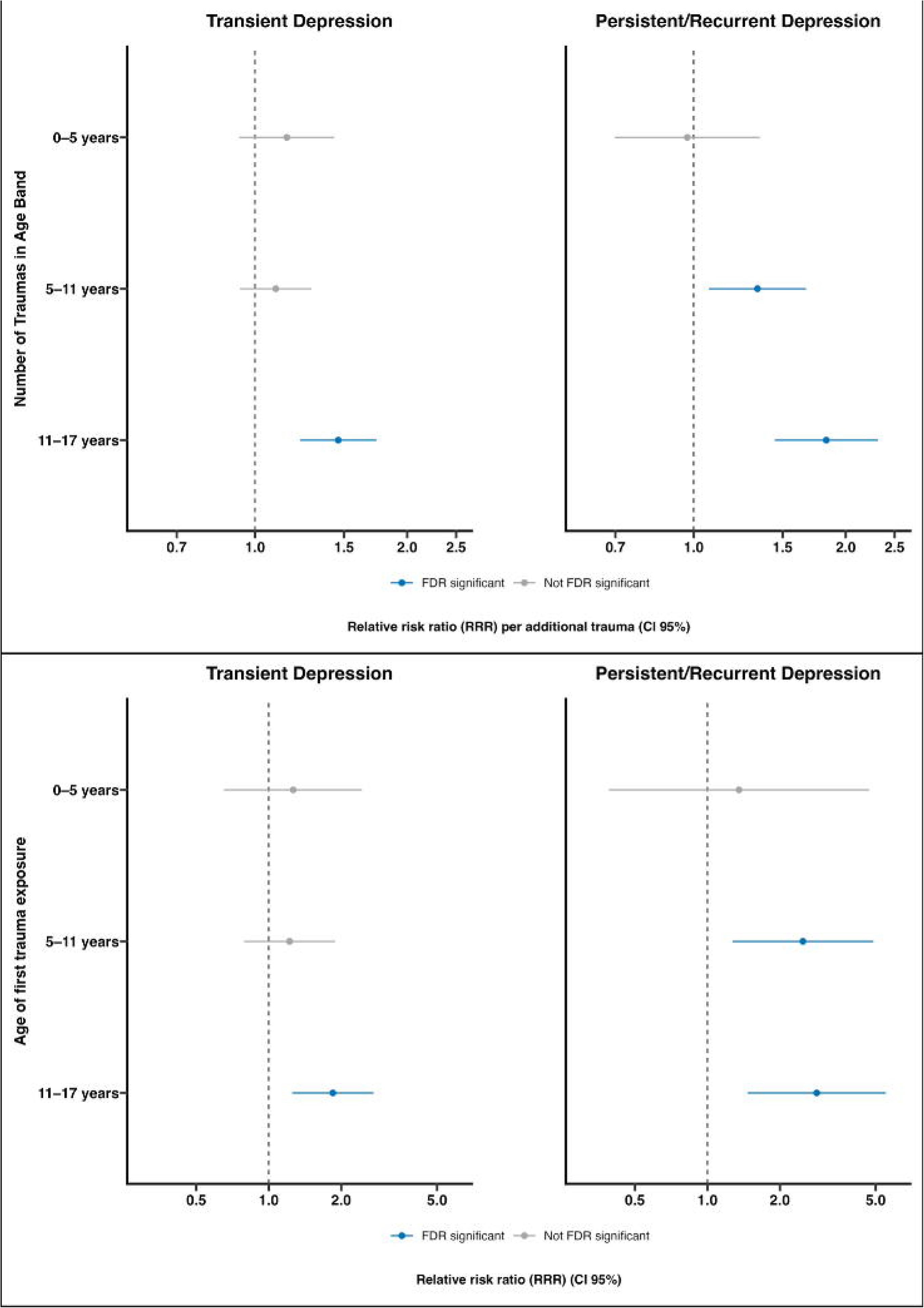
Childhood trauma during different developmental periods and risk of transient depression (one episode), persistent or recurrent depression (two or more episodes) across ages 18, 24 and 30 years. Note: (A) Relative risk ratios (RRRs) and 95% confidence intervals (CIs) from multinomial logistic regression models examining associations between any trauma experienced during early childhood (0-5 yrs), middle childhood (5-11 yrs), and adolescence (11-17 yrs) and Transient and Persistent/Recurrent depression longitudinal groups (Never group as the reference category). (B) RRRs and 95% CIs from multinomial logistic regression models examining associations between the number of traumatic experiences during each developmental period and depression longitudinal group membership. Presented are adjusted models; controlling for sex, ethnicity, maternal age at delivery, maternal education, and maternal social class; see supplementary results for other models. Blue estimates remained significant after false discovery rate (FDR) correction, whereas grey estimates were not FDR significant.

Both any trauma exposure and trauma count during middle childhood (5-11 yrs) were associated with persistent/recurrent depression (RRR = 2.49, 95% CI 1.27-4.90, FDR-P = 0.0119; and RRR = 1.34, 95% CI 1.07-1.67, FDR-P = 0.0151, respectively), with little evidence of associations with transient depression.

Trauma exposure in late childhood/adolescence (11-17 yrs) emerged as the period most strongly associated with later depression. Participants exposed to any trauma during this period had almost three times the risk of persistent/recurrent depression (RRR = 2.85, 95% CI 1.47-5.52, FDR-P = 0.00572), and nearly two-fold higher risk of the transient depression (RRR = 1.85, 95% CI 1.25-2.73, FDR-P = 0.00609), compared to the never depressed group. Similarly, each additional traumatic event experienced between 11-17 yrs was associated with an 83% increase in the risk of persistent/recurrent depression (RRR = 1.83, 95% CI 1.45-2.32, FDR-P = 1.42 × 10⁻⁶) and a 46% increase in the risk of transient depression (RRR = 1.46, 95% CI 1.23-1.74, FDR-P = 5.8 × 10⁻^6^).

To determine whether these findings simply reflected the temporal proximity between trauma exposure (11-17 yrs) and depression measurement (the first CIS-R assessment was at 18 yrs), we next examined the associations between trauma experienced between 11-17 yrs and depression at ages 18, 24 and 30 yrs (**Figure 4**). These results revealed that trauma occurring between 11-17 yrs had similar effects on depression at ages 18, 24 and 30 yrs. Using the continuous trauma count measure, we found that with each additional traumatic event was associated with increased odds of depression at age 18 (OR = 1.59, 95% CI 1.30-1.94, FDR-P = 2.32 x 10^-6^), age 24 (OR = 1.84, 95% CI 1.52-2.22, FDR-P =1.23 x 10^-9^), and age 30 (OR = 1.26, 95% CI 1.02-1.56, FDR-P = 4.64 x 10^-2^). These findings suggest that the stronger associations observed for trauma during adolescence are unlikely to be explained solely by the temporal proximity of trauma exposure to the first depression assessment.

**Figure 4.**
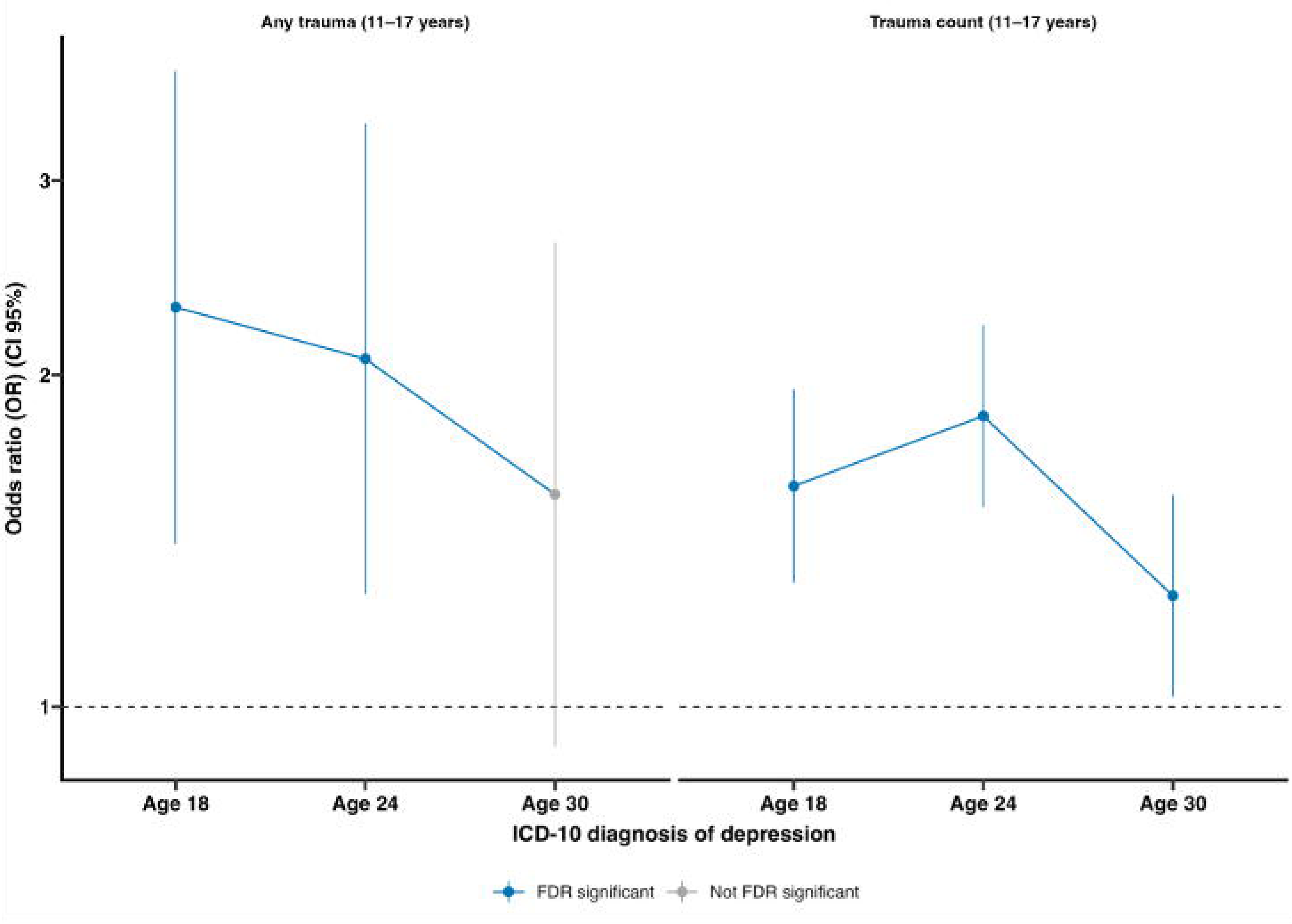
Associations between adolescent trauma and depression at ages 18, 24 and 30 years. Note: Odds ratios (ORs) and 95% confidence intervals (CIs) for associations between trauma experienced at ages 11-17 years and ICD-10 depression assessed using the CIS-R at ages 18, 24 and 30 years. The left panel shows associations for any trauma (exposed versus unexposed), and the right panel shows associations per additional traumatic event. Points represent ORs and vertical bars indicate 95% CIs. Blue estimates remained significant after false discovery rate (FDR) correction, whereas grey estimates were not FDR significant. The dashed horizontal line denotes no association (OR = 1). Models were adjusted for sex, ethnicity, maternal age at delivery, maternal education and maternal social class.

### 4.4 Childhood adversity/trauma is associated with increased circulating IL-6 levels

Childhood adversity/trauma was associated with higher IL-6 concentrations at age 24 after adjustment for potential confounders, although effect sizes were generally modest. Associations were observed for ACE score (β = 0.027, 95% CI 0.002-0.052, FDR-P = 0.036), trauma count (β = 0.050, 95% CI 0.025-0.076, FDR-P = 4.55 × 10⁻⁴), and any trauma (β = 0.100, 95% CI 0.040-0.160, FDR-P = 0.002) (see **Figure 5**, **Supplementary Table 9**).

**Figure 5.**
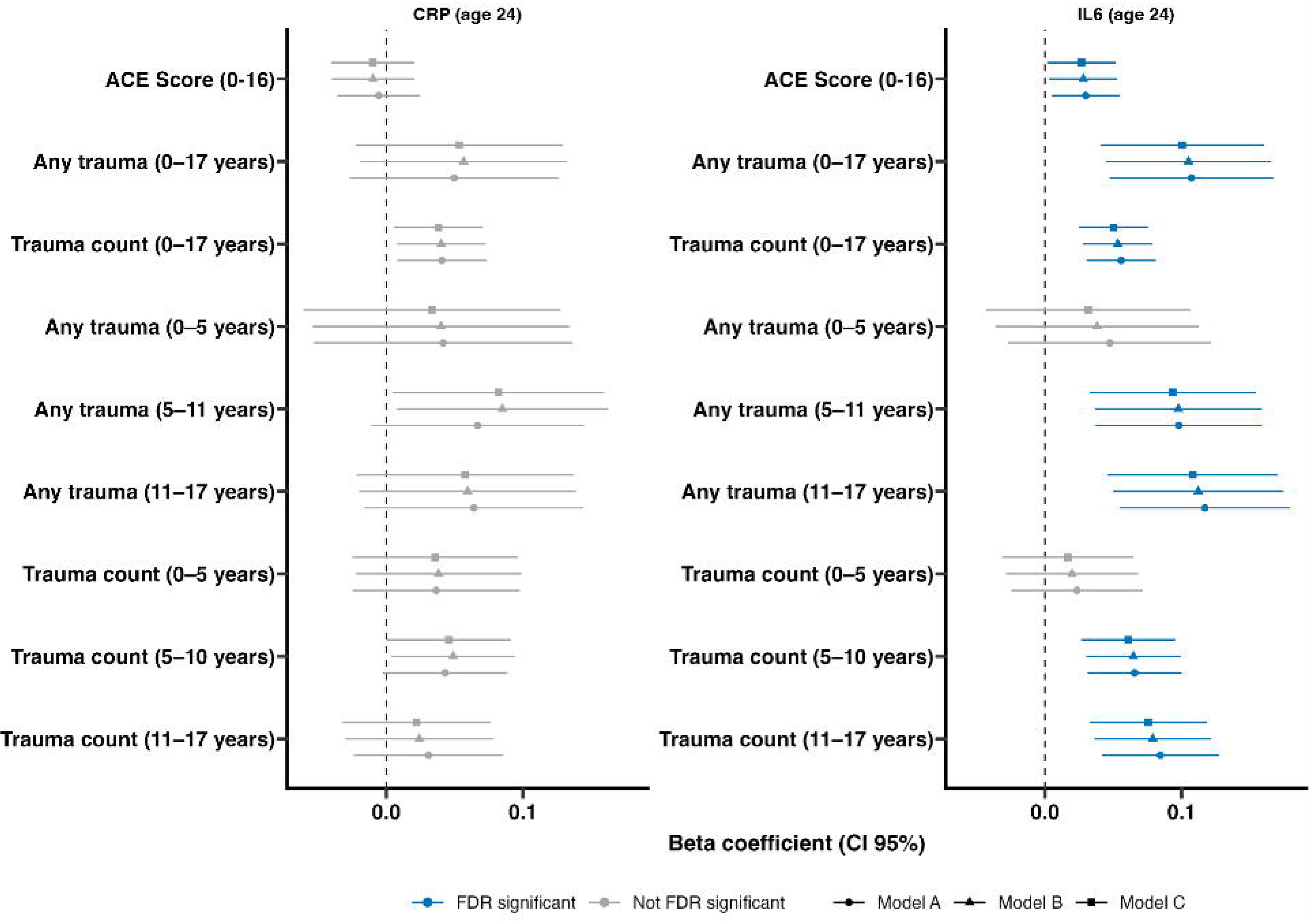
Association of childhood adversity/trauma with circulating CRP and IL-6 levels at age 24 years in the ALSPAC cohort. Note: Beta co-efficient and 95% confidence intervals (CIs) from linear regression models examining associations between ACE score (0-16 yrs), any trauma and trauma count experienced during early childhood (0-5 yrs), middle childhood (5-11 yrs), and adolescence (11-17 yrs), throughout childhood (0-16/17 yrs) and CRP and IL-6 levels at age 24. Models were adjusted sequentially: Model A (circles) was unadjusted; Model B (triangles) was adjusted for sex, ethnicity, maternal age at delivery, maternal education, and maternal social class; and Model C (diamonds) was additionally adjusted for sampling variables (month and time of sample collection, and recency of infection), where these data were available. Blue estimates remained significant after false discovery rate (FDR) correction, whereas grey estimates were not FDR significant.

Timing-specific analyses showed associations with higher IL-6 at age 24 for trauma exposure during middle childhood (5-11 yrs) and adolescence (11-17 yrs). Associations were observed for trauma count during both periods (5-11 years: β = 0.061, 95% CI 0.026-0.096, FDR-P = 7.24 × 10⁻⁴; 11-17 years: β = 0.076, 95% CI 0.033-0.118, FDR-P = 7.25 × 10⁻⁴) and for any trauma (5-11 years: β = 0.093, 95% CI 0.032-0.155, FDR-P = 0.004; 11-17 years: β = 0.108, 95% CI 0.046-0.171, FDR-P = 0.002). Results were similar across models adjusting for biomarker collection covariates and sociodemographic characteristics.

There was little evidence that trauma exposure during early childhood (0-5 yrs), assessed as either trauma count or any trauma, was associated with IL-6 at age 24. Associations between trauma exposure and CRP at age 24 did not survive correction for multiple testing (see **Supplementary Table 9** and **Supplementary Results**).

### 4.5 IL-6 or CRP levels do not mediate the trauma-depression association

Overall, across extensive analyses, there was little evidence that IL-6 or CRP mediated associations between childhood adversity/trauma and ICD-10 depression at age 30 (**Supplementary Table 10**). Across models examining overall trauma exposure, cumulative trauma and exposure during specific childhood periods, estimated proportions mediated ranged from -1.4% to -0.1% for CRP and from 4.4% to 13.2% for IL-6, although confidence intervals were wide and all included zero. Indirect-effect estimates were close to the null (1.00-1.03), with all 95% confidence intervals including 1.

### 4.6 Sensitivity analysis results

Sensitivity analyses generally supported the main findings. Including participants with CRP concentrations >10 mg/L and excluding participants with incomplete depression assessments produced small changes in effect estimates. Analyses restricted to participants with observed covariate data also yielded similar estimates, although some associations crossed the FDR-adjusted threshold. Full results are provided in **Supplementary Results** and **Supplementary Tables 11a-c.**

## 5 Discussion

Persistent or recurrent depression affects approximately 1 in 20 young adults across ages 18, 24 and 30 years. Our findings showed that childhood trauma was consistently associated with depression in young adulthood, with stronger associations for persistent/recurrent depression than for transient depression. Threat-related adversities showed the strongest association with persistent/recurrent depression, and cumulative trauma showed a clear dose-response relationship, suggesting that both the type and burden of childhood adversity are important risk factors for depression into adulthood. Although trauma was associated with higher systemic inflammatory marker levels in later childhood (11-17 yrs), these did not appear to be a major explanatory pathway linking trauma exposure to depression. Incorporating newly available age-30 depression assessments allowed us to extend the study of depression course beyond early adulthood.

Extensive evidence links childhood trauma/adversity with later depression (Flouri et al., 2020; Iob et al., 2022; LeMoult et al., 2020; Nanni et al., 2012; Tan & Mao, 2023). Our findings suggest stronger associations with persistent/recurrent than transient depression, building on evidence that childhood adversity is associated not only with depression onset but also with its chronicity, recurrence and poorer treatment outcomes (Nanni et al., 2012).

Our findings suggest that middle childhood (5-11 yrs) and adolescence (11-17 yrs) may be sensitive periods for adversity-related depression risk. This aligns with evidence linking ACEs at ages 11-17 with depression in early adulthood (Tran et al., 2026). Previous ALSPAC research also linked moderate or high adversity exposure with greater depression risk and severity regardless of timing (Tracy et al., 2019), highlighting the importance of considering both developmental timing and overall adversity burden. However, apparent timing effects may reflect recency or accumulated exposure rather than distinct sensitive periods (Dunn et al., 2018).

We therefore examined whether the adolescent association reflected proximity to the first depression assessment at age 18. Trauma between 11-17 yrs was associated with depression at ages 18 and 24, and trauma count remained associated with depression at age 30, suggesting longer-term relevance beyond proximity to the first outcome assessment, although associations did weaken over time. Notably, a similar timing pattern was observed for inflammation, with trauma exposure in mid to late childhood also showing stronger associations with IL-6 at age 24. Associations between childhood trauma/adversity and CRP were largely attenuated to the null, consistent with findings from previous studies (Crick et al., 2022; Flouri et al., 2020).

Our work highlights that trauma exposure, particularly threat-related adversity and cumulative trauma burden, may be more strongly associated with recurrent patterns of depression than with isolated depressive episodes. The strong associations observed for threat-related adversities are in line with dimensional models of childhood adversity, particularly the threat-deprivation framework proposed by McLaughlin and colleagues, which suggests that experiences involving threat, such as physical, sexual, or emotional abuse, may be especially relevant to socioemotional processing and later psychopathology (McLaughlin et al., 2014; Sheridan & McLaughlin, 2014). Identifying trauma types that are more strongly associated with later mental health may help inform more targeted approaches to prevention and intervention.

Our work adds to existing reports showing that while childhood trauma is associated with higher levels of systemic inflammatory markers such as IL-6 and CRP, these do not fully mediate the trauma-depression relationship (Flouri et al., 2020; Iob et al., 2022). There are several possible explanations for this finding. Raised inflammatory markers do not reflect/capture the aspect of immune response “shaped” or “programmed” by experiencing adversity/trauma. It is also possible that raised inflammatory marker levels are not key mediating pathway linking trauma to depression, but other mechanisms are involved. Hypothalamic-pituitary-adrenal (HPA) axis functioning, which regulates cortisol and stress responsivity (Herman et al., 2016), has been suggested as a possible mediator.

Childhood adversity has been proposed to increase depression risk through allostatic overload of the HPA axis (Wilkinson & Goodyer, 2011), and broader evidence suggests that early adversity can become biologically embedded through interacting changes in stress physiology, immune function, neural development, and epigenetic regulation (Takahashi & Shelton, 2025; Vargovic et al., 2026; Zhou & Ryan, 2023). These processes are highly relevant to depression, as dysregulated cortisol signalling, altered immune activity, and stress sensitisation may increase vulnerability to low mood, anhedonia, fatigue, cognitive symptoms, and impaired emotion regulation (Rousson et al., 2020). Over time, such biological changes may lower resilience to later stressors, helping to explain why childhood trauma had greater association with persistent/recurrent depression than with transient depression. Therefore, the absence of IL-6/CRP mediation in the present study does not rule out biological mechanisms but suggests that a single circulating inflammatory marker may be insufficient to capture the relevant pathways.

Strengths of this study includes the use of ALSPAC, a rich longitudinal cohort spanning childhood, adolescence, and young adulthood. The inclusion of newly available CIS-R data at age 30 allowed depression trajectories to be extended beyond early adulthood, distinguishing persistent/recurrent from transient depression over a longer period than previous ALSPAC studies. The detailed characterisation of childhood adversity allowed examination of cumulative trauma burden, specific trauma types, and developmental timing. The availability of inflammatory biomarker data across childhood allowed trauma-related differences in systemic inflammation to be assessed across development. Multiple imputation, incorporating a range of auxiliary variables, was used to reduce bias from missing data; imputed findings were broadly consistent with complete-case sensitivity analyses, although complete-case estimates were less precise.

Limitations include attrition over time, limited geographic and ethnic diversity, and potential biases arising from self-reported or parent-reported data. Notably, trauma exposure was higher among participants with missing depression repeat assessment data than among those in either observed depression group, suggesting that attrition may have led to underestimation, rather than overestimation, of the association between childhood trauma and later depression.

Prospective assessment of childhood adversity is a key strength. However, most ALSPAC adversity measures before age 8 were parent-reported. Although both parent-and child-reported trauma predict later mental health outcomes (Croft et al., 2019), caregivers may underreport maltreatment (Dunn et al., 2026). While reporting source may have introduced measurement error, the observed associations are unlikely to be entirely attributable to one informant type. Possible underreporting, particularly in early childhood, may have led to exposure misclassification and attenuation of associations with later depression.

In conclusion, our study shows the extent of persistent or recurrent depression between late adolescence and young adulthood between ages 18 and 30 years, which affect approx. 1 in 20 individuals. It also identifies abuse/maltreatment, especially threat adversities experienced during middle and late childhood as potentially important risk factors for persistent depression in young adults. This work also highlights research gaps by highlighting the need for identifying biological mechanisms linking early-life adversity and adult depression.

## Supporting information

Supplemental methods and results

## Acknowledgements

We are extremely grateful to all the families who took part in this study, the midwives for their help in recruiting them, and the whole ALSPAC team, which includes data collection staff, data and administrations staff, technical managers and the technical staff with the Bristol Bioresource Laboratory, based within the University of Bristol.

## Data availability

The informed consent obtained from ALSPAC (Avon Longitudinal Study of Parents and Children) participants does not allow the data to be made available through any third party maintained public repository. Supporting data are available from ALSPAC on request under the approved proposal number, B4864. Full instructions for applying for data access can be found here: http://www.bristol.ac.uk/alspac/researchers/access/. The ALSPAC study website contains details of all available data (http://www.bristol.ac.uk/alspac/researchers/our-data/).

## 7 Supplementary Material

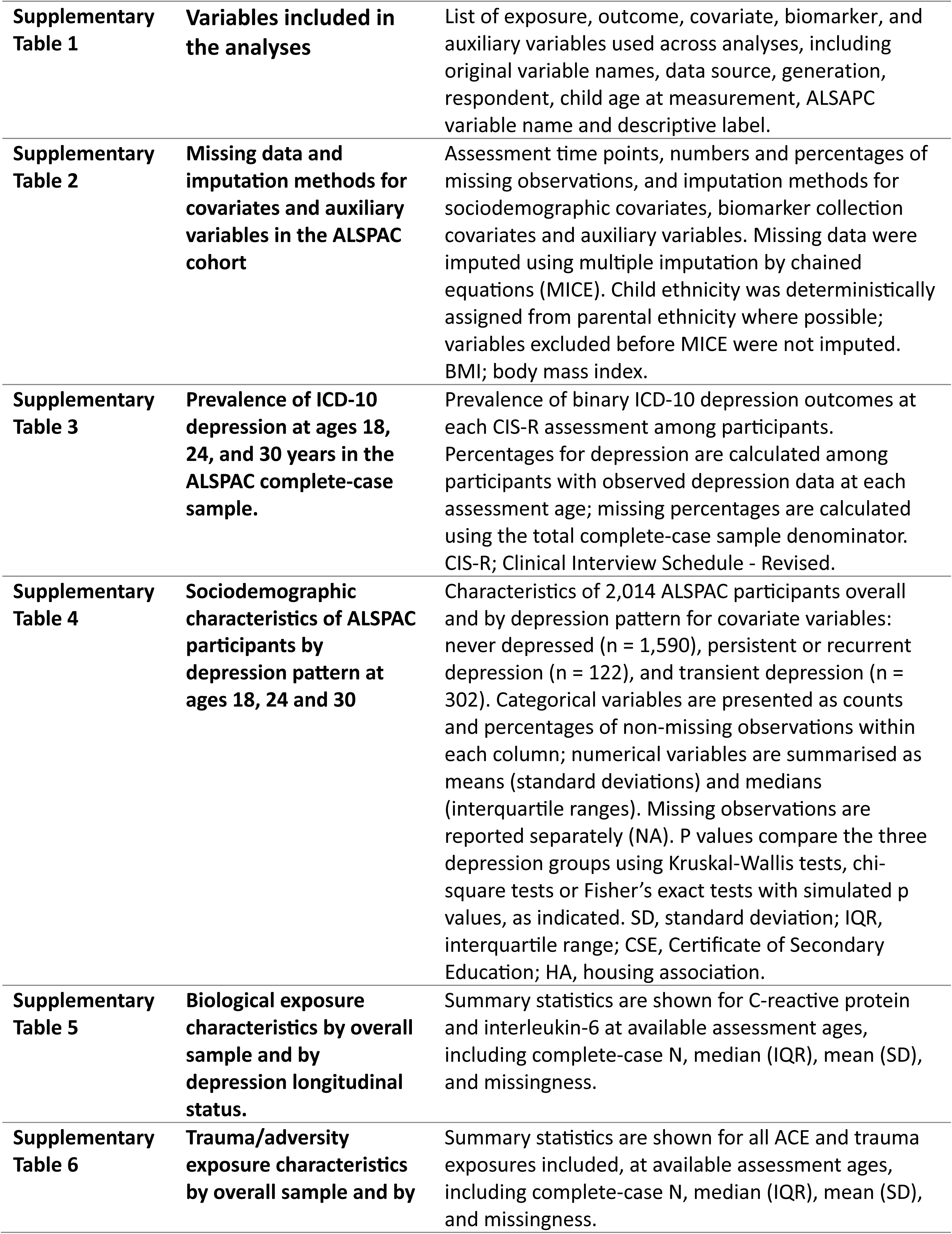
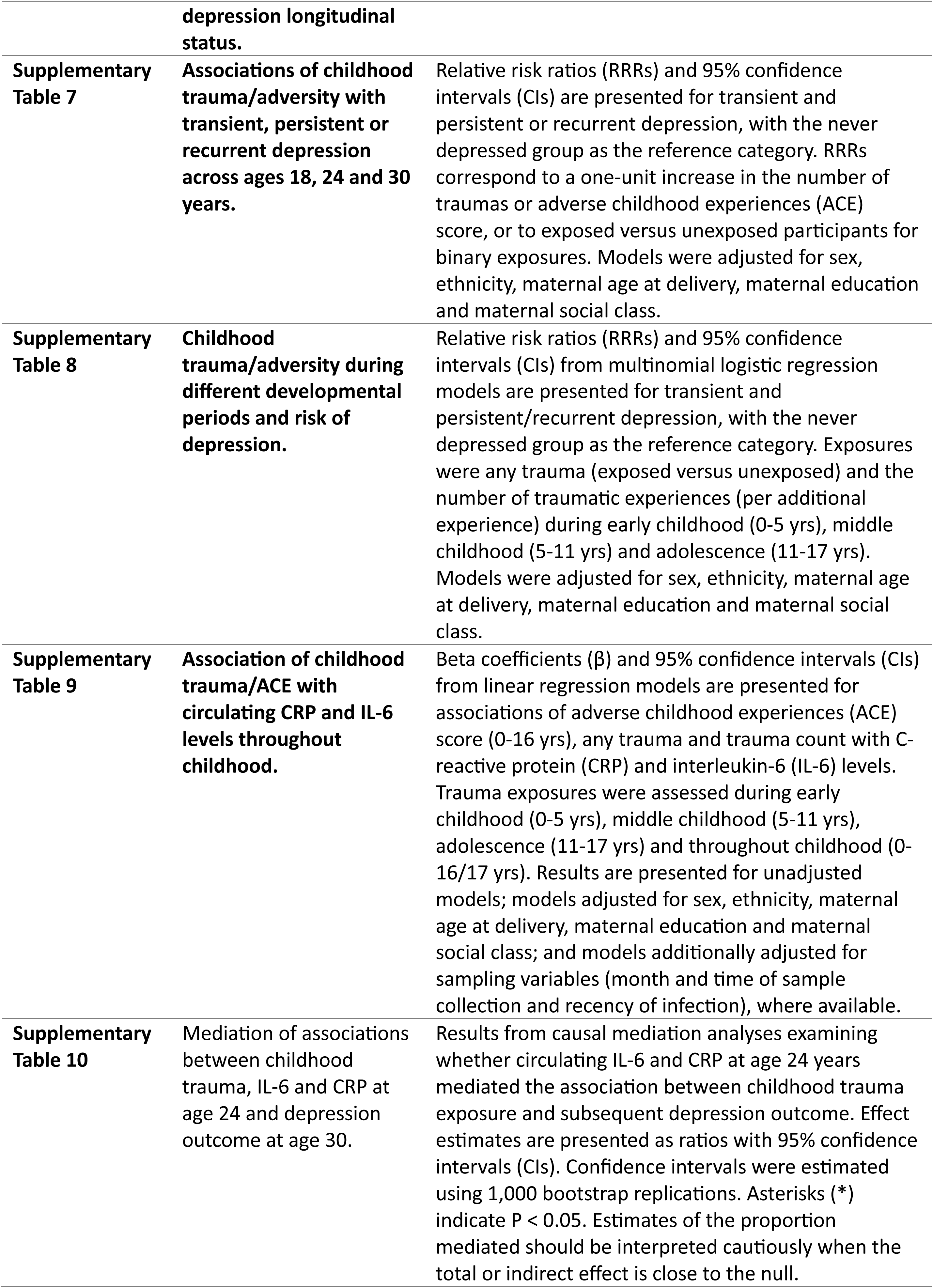
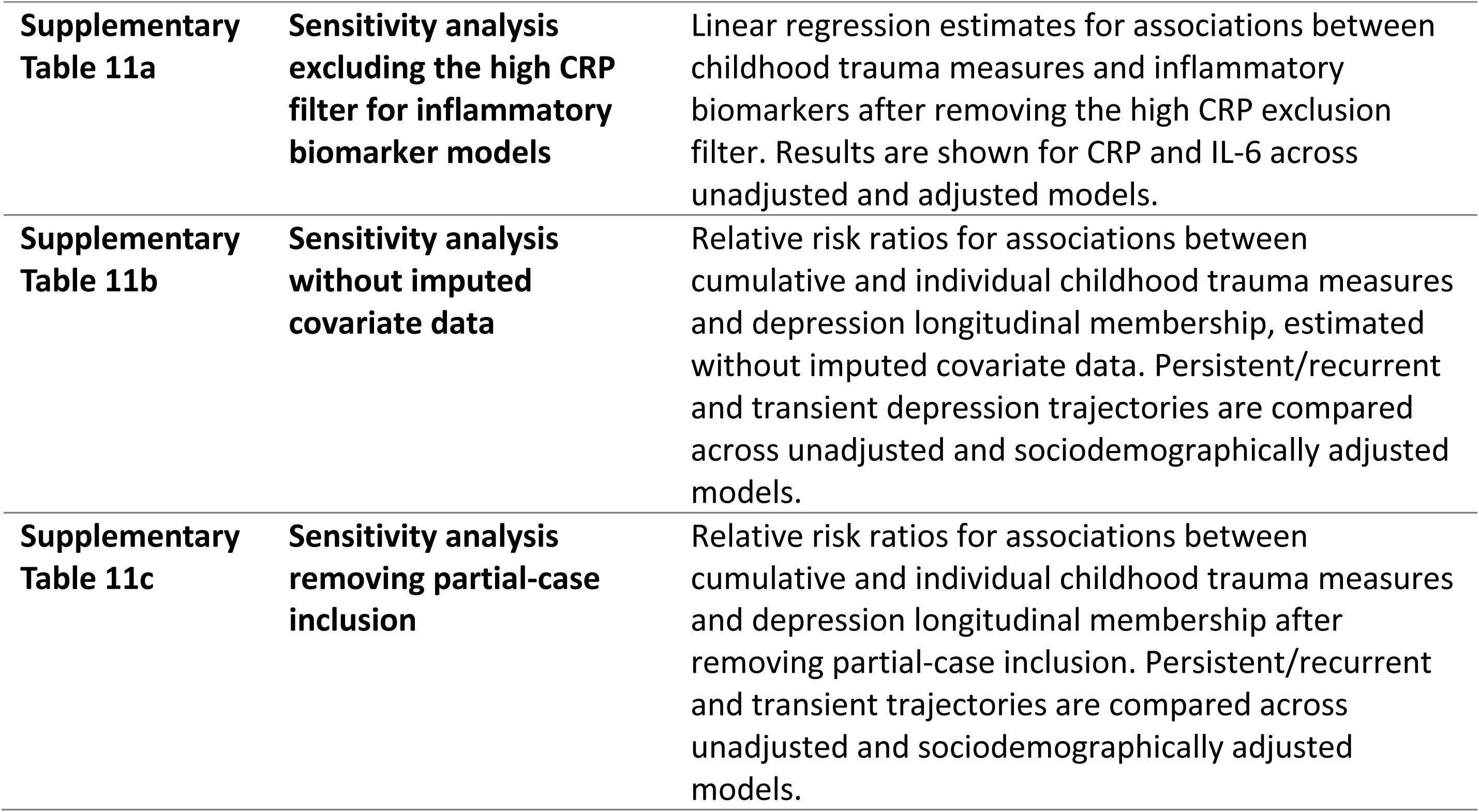

|  |  |  |
|---|---|---|
| **Supplementary Table 1** | **Variables included in the analyses** | List of exposure, outcome, covariate, biomarker, and auxiliary variables used across analyses, including original variable names, data source, generation, respondent, child age at measurement, ALSAPC variable name and descriptive label. |
| **Supplementary Table 2** | **Missing data and imputation methods for covariates and auxiliary variables in the ALSPAC cohort** | Assessment time points, numbers and percentages of missing observations, and imputation methods for sociodemographic covariates, biomarker collection covariates and auxiliary variables. Missing data were imputed using multiple imputation by chained equations (MICE). Child ethnicity was deterministically assigned from parental ethnicity where possible; variables excluded before MICE were not imputed. BMI; body mass index. |
| **Supplementary Table 3** | **Prevalence of ICD-10 depression at ages 18, 24, and 30 years in the ALSPAC complete-case sample.** | Prevalence of binary ICD-10 depression outcomes at each CIS-R assessment among participants. Percentages for depression are calculated among participants with observed depression data at each assessment age; missing percentages are calculated using the total complete-case sample denominator. CIS-R; Clinical Interview Schedule - Revised. |
| **Supplementary Table 4** | **Sociodemographic characteristics of ALSPAC participants by depression pattern at ages 18, 24 and 30** | Characteristics of 2,014 ALSPAC participants overall and by depression pattern for covariate variables: never depressed (n = 1,590), persistent or recurrent depression (n = 122), and transient depression (n = 302). Categorical variables are presented as counts and percentages of non-missing observations within each column; numerical variables are summarised as means (standard deviations) and medians (interquartile ranges). Missing observations are reported separately (NA). P values compare the three depression groups using Kruskal-Wallis tests, chi-square tests or Fisher’s exact tests with simulated p values, as indicated. SD, standard deviation; IQR, interquartile range; CSE, Certificate of Secondary Education; HA, housing association. |
| **Supplementary Table 5** | **Biological exposure characteristics by overall sample and by depression longitudinal status.** | Summary statistics are shown for C-reactive protein and interleukin-6 at available assessment ages, including complete-case N, median (IQR), mean (SD), and missingness. |
| **Supplementary Table 6** | **Trauma/adversity exposure characteristics by overall sample and by depression longitudinal status.** | Summary statistics are shown for all ACE and trauma exposures included, at available assessment ages, including complete-case N, median (IQR), mean (SD), and missingness. |
| **Supplementary Table 7** | **Associations of childhood trauma/adversity with transient, persistent or recurrent depression across ages 18, 24 and 30 years.** | Relative risk ratios (RRRs) and 95% confidence intervals (CIs) are presented for transient and persistent or recurrent depression, with the never depressed group as the reference category. RRRs correspond to a one-unit increase in the number of traumas or adverse childhood experiences (ACE) score, or to exposed versus unexposed participants for binary exposures. Models were adjusted for sex, ethnicity, maternal age at delivery, maternal education and maternal social class. |
| **Supplementary Table 8** | **Childhood trauma/adversity during different developmental periods and risk of depression.** | Relative risk ratios (RRRs) and 95% confidence intervals (CIs) from multinomial logistic regression models are presented for transient and persistent/recurrent depression, with the never depressed group as the reference category. Exposures were any trauma (exposed versus unexposed) and the number of traumatic experiences (per additional experience) during early childhood (0-5 yrs), middle childhood (5-11 yrs) and adolescence (11-17 yrs). Models were adjusted for sex, ethnicity, maternal age at delivery, maternal education and maternal social class. |
| **Supplementary Table 9** | **Association of childhood trauma/ACE with circulating CRP and IL-6 levels throughout childhood.** | Beta coefficients (β) and 95% confidence intervals (CIs) from linear regression models are presented for associations of adverse childhood experiences (ACE) score (0-16 yrs), any trauma and trauma count with C-reactive protein (CRP) and interleukin-6 (IL-6) levels. Trauma exposures were assessed during early childhood (0-5 yrs), middle childhood (5-11 yrs), adolescence (11-17 yrs) and throughout childhood (0-16/17 yrs). Results are presented for unadjusted models; models adjusted for sex, ethnicity, maternal age at delivery, maternal education and maternal social class; and models additionally adjusted for sampling variables (month and time of sample collection and recency of infection), where available. |
| **Supplementary Table 10** | Mediation of associations between childhood trauma, IL-6 and CRP at age 24 and depression outcome at age 30. | Results from causal mediation analyses examining whether circulating IL-6 and CRP at age 24 years mediated the association between childhood trauma exposure and subsequent depression outcome. Effect estimates are presented as ratios with 95% confidence intervals (CIs). Confidence intervals were estimated using 1,000 bootstrap replications. Asterisks (*) indicate P < 0.05. Estimates of the proportion mediated should be interpreted cautiously when the total or indirect effect is close to the null. |
| **Supplementary Table 11a** | **Sensitivity analysis excluding the high CRP filter for inflammatory biomarker models** | Linear regression estimates for associations between childhood trauma measures and inflammatory biomarkers after removing the high CRP exclusion filter. Results are shown for CRP and IL-6 across unadjusted and adjusted models. |
| **Supplementary Table 11b** | **Sensitivity analysis without imputed covariate data** | Relative risk ratios for associations between cumulative and individual childhood trauma measures and depression longitudinal membership, estimated without imputed covariate data. Persistent/recurrent and transient depression trajectories are compared across unadjusted and sociodemographically adjusted models. |
| **Supplementary Table 11c** | **Sensitivity analysis removing partial-case inclusion** | Relative risk ratios for associations between cumulative and individual childhood trauma measures and depression longitudinal membership after removing partial-case inclusion. Persistent/recurrent and transient trajectories are compared across unadjusted and sociodemographically adjusted models. |

**Supplementary Methods 1 - Inflammatory Biomarkers collection methods**

**Supplementary Methods 2 - Depression longitudinal classification**

**Supplementary Methods 3 - Potential confounders and adjusted models**

**Supplementary Methods 4 - Missing Data and Multiple Imputation**

**Supplementary Methods 5 - Sensitivity analysis**

