## Supplemental methods and results for "Persistent or recurrent depression in young adults: associations with systemic inflammation and early life adversity"

#### Supplementary Methods and Results

Persistent or recurrent depression in young adults: associations with systemic inflammation and early life adversity.

### Supplementary Methods

#### 1 Inflammatory biomarkers collection and measurement

##### 1.1 CRP at ages 9, 16, 18 and 24 years

High-sensitivity CRP was measured in blood samples collected during childhood, adolescence and young adulthood. Samples collected at age 9 years were obtained from non-fasting participants, whereas participants attending the adolescent and age-24 assessments were instructed to fast overnight or for at least six hours before blood collection. Samples were centrifuged and stored at −80°C before analysis.

At each assessment, CRP concentrations >10 mg/L were set to missing for that timepoint only to reduce the influence of elevations potentially reflecting acute infection or other inflammatory conditions (Pearson et al. 2003). The remaining values were standardised to Z-scores separately at each assessment before analysis.

##### 1.2 IL-6 at age 9 years

IL-6 was measured in blood samples collected from non-fasting participants attending the Focus@9 clinic. Samples were centrifuged immediately after collection and stored at −80°C. IL-6 concentrations were measured using an enzyme-linked immunosorbent assay (ELISA; R&D Systems) and expressed in pg/mL.

##### 1.3 IL-6 at age 24 years

Plasma samples were obtained from 3,027 fasting offspring attending the Focus@24 clinic. IL-6 was measured in heparin plasma using the Olink Target 96 Inflammation panel, which uses proximity extension assay technology. Measurements were expressed as normalised protein expression (NPX) values on a $\log_2$ scale, representing relative protein abundance rather than absolute concentrations. Detailed sample processing, assay and quality control procedures have been described previously (Goulding et al. 2024).

#### 2 Depression trajectory classification

##### 2.1 Never, Persistent, Recurrent and Transient

2014 participants had data on all three depression outcomes at ages 18, 24 and 30. Each timepoint is coded as either 0 = no depression; 1 = depression present; or, NA. This creates 8 options over the 3 time points.

1. 0 0 0 (baseline in modelling)
2. 0 0 1
3. 0 1 0
4. 0 1 1
5. 1 0 0
6. 1 0 1

7. 1 1 0
8. 1 1 1

| Never (baseline) | Persistent<br>(two consecutive timepoints)<br>Recurrent<br>(two non-consecutive timepoints) | Transient<br>(one timepoint) |
|---|---|---|
| 0 0 0 | 011 | 100 |
| | 110 | 010 |
| | 111 | 001 |
| | 101 | |

#### 2.2 Partial-case inclusion using incomplete cases

In addition to complete cases, the primary analysis included an additional 48 participants who had missing data at one timepoint, where their trajectory classification would not change regardless of the missing value.

Additional cases included that are logically invariant:

| Observed pattern | Missing wave | Always classifies as |
|---|---|---|
| NA 1 1 | 18 | Persistent/Recurrent |
| 1 NA 1 | 24 | Persistent/Recurrent |
| 1 1 NA | 30 | Persistent/Recurrent |

A sensitivity analysis was also undertaken using complete cases only.

### 3 Potential confounders and adjusted models

#### 3.1 Main potential confounders

Sex. Sex of child was obtained from birth notification [kz021]. NB: Participants with missing sex (n = 576) were excluded from sex-adjusted analyses.

Maternal education (at 32 weeks gestation). Mother's highest educational qualification is a derived variable in ALSPAC which includes the following categories: Degree, A-level, O-level, Vocational, CSE/none. Variable C645A used here is a recode of variable C645. C645A puts all mothers who left education questions blank to “CSE/none”, under the assumption that mothers with no educational qualifications would leave this question blank [C645A].

Maternal occupation (at 32 weeks gestation). Maternal occupation is a set of derived variables in ALSPAC which are based on the Office of Population Censuses and Surveys (OPCS) job codes. It includes the following categories: professional, intermediate, skilled (non-manual), skilled (manual), partly skilled, unskilled, and armed forces. Due to a small number of individuals the armed forces group (N < 4), these individuals were removed. The maternal occupation variable is used as a proxy for maternal socioeconomic position (SES) [C_SOC2000_M].

Child ethnic background. Child ethnic background was reported by the mother during pregnancy [C804].

Maternal age at delivery. Maternal age at delivery is an ALSPAC-derived grouped variable based on the mother’s age at the time of the child’s birth [MZ028B].

#### 3.2 Biological potential confounders

Season of sample taken (at age 9). Season of sample collection at the age 9 clinic was derived from the month of clinic attendance. Month of attendance was recorded as part of the child clinic visit and collapsed into four seasons: Winter (December–February), Spring (March–May), Summer (June–August), and Autumn (September–November). Responses coded as “Did not do samples” were recoded to missing [F9001].

Season of sample taken (at age 24). Season of sample collection at the age 24 clinic was derived from the month of clinic attendance. Month of attendance was recorded as part of the child clinic visit and collapsed into four seasons: Winter (December–February), Spring (March–May), Summer (June–August), and Autumn (September–November). Responses coded as “Did not do samples” were recoded to missing [FKAR0040].

Hour of day of sample taken (at age 9). Hour of day of sample collection at the age 9 clinic was recorded as the sample session start time (hours) during the clinic visit. This variable reflects the time of day at which biological samples were collected. Hour of attendance was recorded as part of the child clinic visit and collapsed into morning (hours before 12) and afternoon (hours after 12. Sample hour responses coded as “Did not do samples” were recoded to missing [F9SA002A].

Hour of day of sample taken (at age 24). Hour of day of sample collection at the age 24 clinic was recorded as the sample session start time (hours) during the clinic visit. This variable reflects the time of day at which biological samples were collected. Hour of attendance was recorded as part of the child clinic visit and collapsed into morning (hours before 12) and afternoon (hours after 12. Sample hour responses coded as “Did not do samples” were recoded to missing [FKCO0010].

Infection - no. of days before visit (age 9). This variable indicates whether the child had any current infection at the time of the clinic visit, irrespective of infection type. Responses coded as “Did not do samples” were recoded as missing [F9SA011]. Internal consistency checks showed full agreement between infection presence and timing variables, with a small proportion of missing timing data among participants reporting infection. NB: there is no comparable infection days before visit for age 24 clinc.

#### 3.3 Model set ups

| Analysis | Model setup | Confounders | Outcome model |
|---|---|---|---|
| Analysis 1: psychosocial adversity and depression trajectory | Model A | None | Multinomial regression, depression trajectory reference group = Never |
| Analysis 1: psychosocial adversity and depression trajectory | Model B | Sex, Ethnic (child), Mother age at delivery (grouped), Mothers highest educational qualification, Maternal social class | Multinomial regression, depression trajectory reference group = Never |
| Analysis 2: developmental timing of trauma and depression | Model A | None | Multinomial regression for depression trajectory; logistic regression for age-specific depression sensitivity outcomes |
| Analysis 2: developmental timing of trauma and depression | Model B | Sex, Ethnic group (child), Mother age at delivery (grouped), Mothers highest educational qualification, Maternal social class | Multinomial regression for depression trajectory; logistic regression for age-specific depression sensitivity outcomes |
| Analysis 3: psychosocial adversity and inflammatory biomarkers, age 9 | Model A | None | Linear regression |
| Analysis 3: psychosocial adversity and inflammatory biomarkers, age 9 | Model B | Month of clinic visit (age 9), Sample collection start time (age 9), Days since infection before clinic visit (age 9) | Linear regression |
| Analysis 3: psychosocial adversity and inflammatory biomarkers, age 9 | Model C | Month of clinic visit (age 9), Sample collection start time (age 9), Days since infection before clinic visit (age 9), Sex, Mothers highest educational qualification, Maternal social class | Linear regression |
| Analysis 3: psychosocial | Model A | None | Linear regression |

| | | | |
|---|---|---|---|
| adversity and inflammatory biomarkers, age 24 | | | |
| Analysis 3: psychosocial adversity and inflammatory biomarkers, age 24 | Model B | Month of clinic visit (age 24), Sample collection start time (age 24) | Linear regression |
| Analysis 3: psychosocial adversity and inflammatory biomarkers, age 24 | Model C | Month of clinic visit (age 24), Sample collection start time (age 24), Sex, Ethnic group (child), Mother age at delivery (grouped), Mothers highest educational qualification, Maternal social class | Linear regression |
| Analysis 4: IL-6 mediation pathway, age 9 mediator | Fully adjusted mediation model | Month of clinic visit (age 9), Sample collection start time (age 9), Days since infection before clinic visit (age 9), Sex, Ethnic group (child), Mother age at delivery (grouped), Mothers highest educational qualification, Maternal social class. BMI as intermediate cofounder. | CMAverse mediation models for depression trajectory or age-18 depression |
| Analysis 4: IL-6 mediation pathway, age 24 mediator | Fully adjusted mediation model | Month of clinic visit (age 24), Sample collection start time (age 24), Sex, Ethnic group (child), Mother age at delivery (grouped), Mothers highest educational qualification, Maternal social class. BMI as intermediate cofounder. | CMAverse mediation models for depression trajectory or age-30 depression |

### 4 Missing Data and Multiple Imputation

#### 4.1 Missingness

Missingness in covariates and auxiliary variables before imputation is reported in Supplementary Table 2. Missingness was calculated after data cleaning and before multiple imputation.

#### 4.2 Multiple Imputation Methods

Missing covariate data were handled using multiple imputation by chained equations (MICE), implemented in R using the `mice` package. Multiple imputation was conducted for covariates and auxiliary variables only; outcome variables were not included in the imputation models. Twenty imputed datasets were generated using fully conditional specification over 20 iterations.

Variables were preprocessed before imputation according to their measurement type. Continuous variables were treated as numeric, binary variables were encoded as two-level factors, ordinal variables were specified as ordered factors, and nominal categorical variables were treated as unordered factors.

The imputation method was then matched to variable type: continuous variables were imputed using predictive mean matching, binary variables using logistic regression, ordinal variables using proportional odds ordinal logistic regression, and nominal categorical variables using multinomial logistic regression.

The imputation model included the analysis covariates together with auxiliary variables selected a priori because of their substantive relevance and plausible associations with either covariate missingness or the incomplete variables.

These auxiliary variables included related anthropometric, sociodemographic, household, maternal, and child health measures from earlier and later assessment waves. Supplementary Table S5 lists the variables included in the imputation setup, their pre-imputation missingness, and the regression model used for each imputed variable.

The participant identifier was not imputed and was excluded from the predictor matrix. The predictor matrix was constructed using `mice::quickpred()`, with a minimum correlation threshold of 0.05 and a minimum proportion of usable cases of 0.25, so that predictors were retained where there was sufficient pairwise information. Sex, maternal age at delivery, child ethnicity, maternal ethnicity, and partner ethnicity were forced into the predictor matrix where available. Variables with very limited observed information were not used as predictors or imputation targets.

Three variables were excluded before creating the MICE object because of very high missingness or limited observed information: days since infection before the age-9 clinic visit, and month of revisit to the age-9 clinic. Child ethnicity was deterministically completed where possible using the observed parental ethnicity rule and was then not imputed by MICE. Maternal and partner ethnicity were complete in the cleaned covariate data and were used as predictors but were not imputed.

Analyses using the imputed covariates were run across the 20 imputed datasets and combined using standard multiple-imputation pooling procedures to account for uncertainty in the imputed values.

### 5 Sensitivity analysis methods

#### 5.1.i Sensitivity analysis including participants with CRP greater than 10 mg/L.

We repeated all biomarker analysis, including participants with C-reactive protein (CRP) concentrations greater than 10 mg/L, who were excluded from the primary biomarker analyses to reduce the influence of possible acute infection or inflammation.

#### 5.1.ii Sensitivity analysis removing multiple imputation of missing covariate data.

We repeated analysis using complete observed covariate data only, excluding imputed covariate values. In these models, participants were retained only where the relevant exposure, outcome, and covariate data were observed for the specific model being fitted. This assessed whether findings were sensitive to the use of multiple imputation for missing covariate data.

#### Sensitivity analysis removing partial-case inclusion of depression trajectories.

We repeated analyses involving depression longitudinal outcomes using the stricter longitudinal classification that excluded participants whose persistent/recurrent classification depended on partial-case inclusion. This sensitivity outcome set removed participants with two observed depressive episodes and one missing depression assessment, retaining only participants complete data at all 3 time points. The resulting analyses used the same model specifications as the primary analyses.

### Supplementary Results

#### 1 Exposures and depression

Inflammatory marker distributions were broadly similar across depression longitudinal groups (**Error! Reference source not found.**), although values tended to be slightly higher among participants in the persistent/recurrent group at later ages. At age 9, median CRP and IL-6 levels were comparable across groups. By age 24, the persistent/recurrent group had higher median IL-6 [3.56 NPX log2, IQR 3.09-4.33] and CRP [1.10 mg/L, IQR 0.42-3.49] than the never group [IL-6: 3.28, IQR 2.92-3.73; CRP: 0.80 mg/L, IQR 0.37-1.91]. Missingness was substantial for inflammatory biomarkers at all time points, including among participants without depression diagnosis (**Error! Reference source not found.**).

Psychosocial exposure characteristics differed descriptively across depression groups. Any reported trauma between ages 0 and 17, and other trauma related exposures, were more frequent among participants with persistent/recurrent (55.2%) or transient depression (65.8%) than among those who were never depressed, however 42.8% of those who were never depressed still experienced trauma during childhood (**Error! Reference source not found.**).

#### 2 Association of childhood trauma/ACE with circulating systemic inflammation.

A)

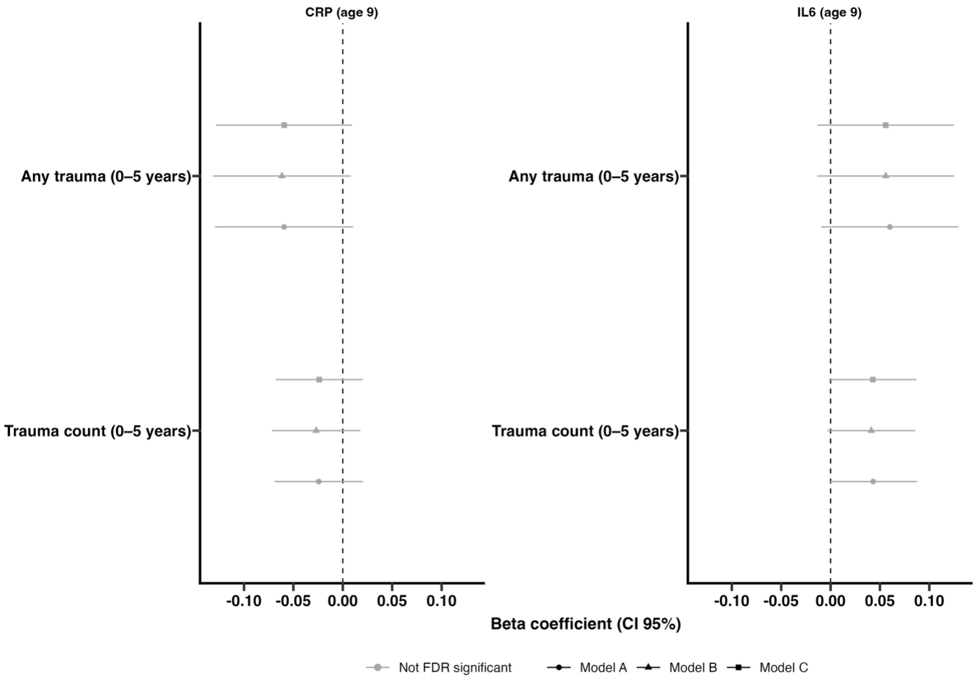
CRP (age 9)
IL6 (age 9)
Any trauma (0–5 years)
Trauma count (0–5 years)
-0.10 -0.05 0.00 0.05 0.10
Beta coefficient (CI 95%)
Not FDR significant
Model A
Model B
Model C

B)

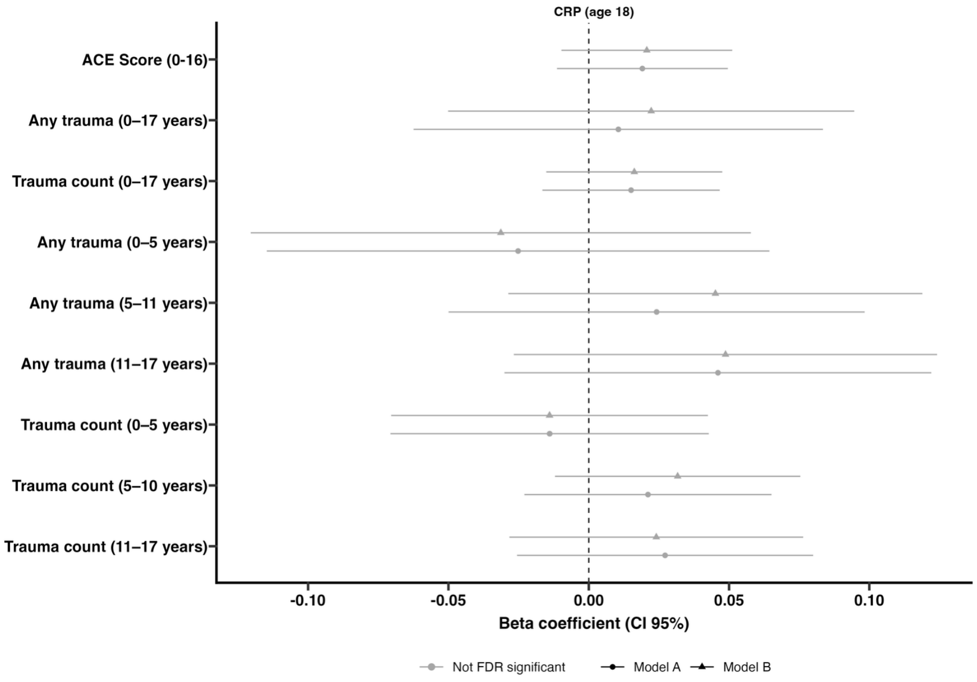
CRP (age 18)
ACE Score (0-16)
Any trauma (0–17 years)
Trauma count (0–17 years)
Any trauma (0–5 years)
Any trauma (5–11 years)
Any trauma (11–17 years)
Trauma count (0–5 years)
Trauma count (5–10 years)
Trauma count (11–17 years)
-0.10 -0.05 0.00 0.05 0.10
Beta coefficient (CI 95%)
Not FDR significant
Model A
Model B

C)

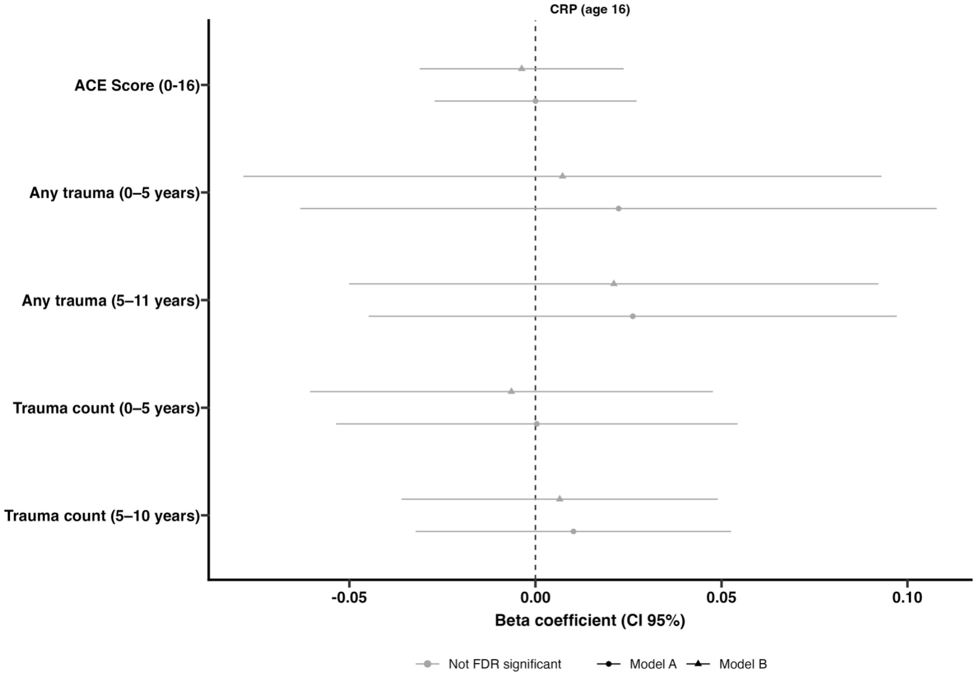

**Supplementary Figure 1. Associations between childhood trauma and IL6/CRP at 9, 16 and 18 yrs.** Beta co-efficient and 95% confidence intervals (CIs) from linear regression models examining associations between ACE score (0-16 yrs), any trauma and trauma count experienced during early childhood (0-5 years), middle childhood (5-11 yrs), and adolescence (11-17 yrs), throughout childhood (0-16/17 yrs) and Transient and Persistent/Recurrent depression (Never as the reference category). Circles represent unadjusted models; triangles represent adjusted models, controlling for sex, ethnicity, maternal age at delivery, maternal education, and maternal social class; diamonds represent also controlling for sampling variables (month and time of sample taken, infection recency) (included where data were available). Blue estimates remained significant after false discovery rate (FDR) correction, whereas grey estimates were not FDR significant.

### 3 Mediation results

As trauma exposure was associated with both subsequent depression and increased inflammatory markers, we next investigated whether circulating IL-6 and CRP at age 24 mediated associations between childhood trauma and later depression. Causal mediation analyses were conducted using a counterfactual framework. To minimise multiple testing, analyses focused on trauma-inflammation associations that remained statistically significant in the previous analyses, while also including the developmental windows most strongly associated with later depression, particularly trauma exposure between ages 5-11 and 11-17 years. Results are presented in **Supplementary Table 10.**

There was no evidence that IL-6 or CRP at age 24 mediated associations between childhood trauma and subsequent depression at age 30. For cumulative exposure measures, total effects indicated that ACE score (ages 0–16 years), any trauma, and trauma count (ages 0–17 years) were associated with higher odds of later depression (ORs = 1.19–2.04; P = 0.002–0.041). However, indirect effects through IL-6 or

CRP were close to the null (ORs = 1.00–1.02; P = 0.192–0.959), and all corresponding 95% confidence intervals included 1. Direct effects were similar in magnitude to total effects (ORs = 1.20–2.04; P = 0.002–0.039), providing no evidence of mediation through either inflammatory marker at age 24.

There was also no evidence of mediation in the timing-specific exposure windows. For trauma count between ages 5–11 years, total effects were associated with depression in both the CRP and IL-6 mediation models, but indirect effects were close to the null (CRP: OR = 1.00, 95% CI 0.98–1.01, P = 0.665; IL-6: OR = 1.02, 95% CI 0.99–1.05, P = 0.144). Any trauma between ages 5–11 years also showed no evidence of mediation by IL-6 (indirect effect: OR = 1.02, 95% CI 0.99–1.06, P = 0.146). For adolescent trauma exposure between ages 11–17 years, indirect effects through IL-6 were again small, with confidence intervals including the null for both any trauma (OR = 1.03, 95% CI 0.99–1.07, P = 0.163) and trauma count (OR = 1.02, 95% CI 0.99–1.06, P = 0.214).

Overall, these findings provide further evidence trauma is associated with higher odds/risk of depression at age 30, but there is little evidence that CRP or IL-6 at age 24 mediates that association, including for the middle childhood and adolescent exposure windows in which trauma-depression associations were strongest.

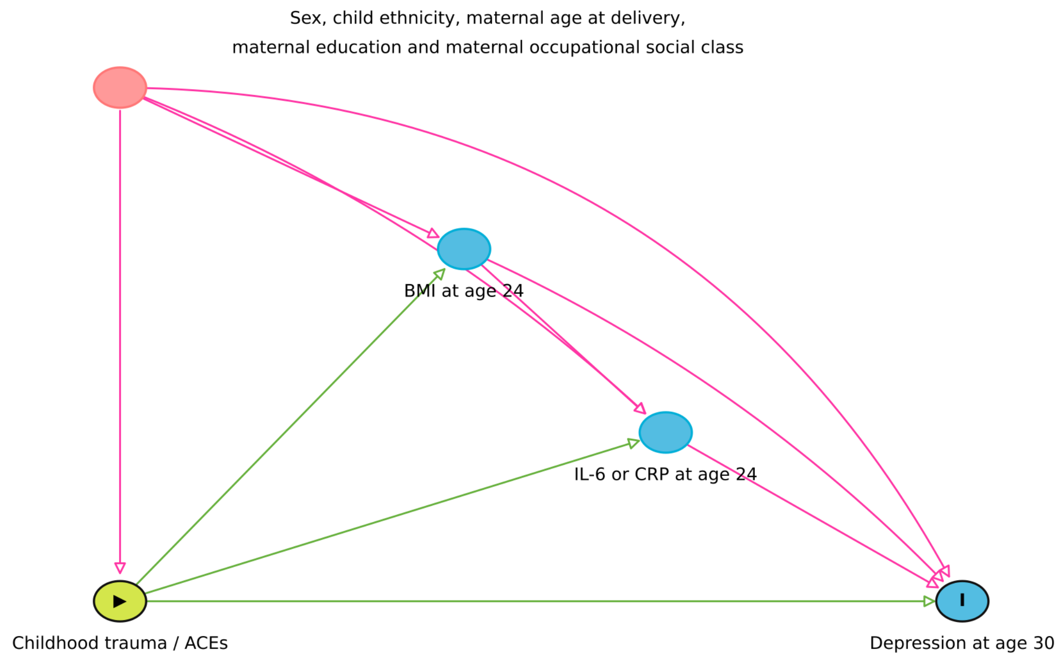

**Supplementary Figure 2. Directed Acyclic Graph (DAG) of Mediation Analysis.** Childhood trauma is modelled as the exposure, IL-6 as a potential mediator, and depression as the outcome. Covariates including child sex, ethnicity, maternal age at delivery, maternal education, and maternal occupation are included as potential confounders of the trauma-inflammation, trauma-depression, and inflammation-depression associations.

### 4 Sensitivity analysis results

Sensitivity analyses showed that the overall pattern of findings was broadly consistent with the main analyses, although some results were sensitive to analytic decisions. For each sensitivity analysis, estimates were compared with the primary analyses in terms of direction, magnitude, confidence intervals, nominal and FDR-adjusted statistical significance, and sample size. Comparator tables were generated to identify results where conclusions changed materially.

#### 4.1.i Results of sensitivity analysis of including participants with high CRP

Participants with high CRP concentrations (>10 mg/L), suggestive of acute infection or inflammation, were excluded from the primary CRP and IL-6 analyses, consistent with standard practice in epidemiological studies of low-grade systemic inflammation (Pearson et al. 2003). Therefore, a sensitivity analysis was conducted which included these participants. This resulted in an increase of 53 participants across comparisons; which had a small effect on the results. However, the median absolute change in beta was small, 0.018, and only 2 sensitivity estimates fell outside the corresponding main-analysis 95% confidence interval (see Supplementary Table 11a for results).

#### 4.1.ii Results sensitivity analysis of excluding imputed covariates

When analyses were repeated using observed covariate data only, rather than imputed covariates, effect estimates were generally very similar to the main analyses (Supplementary Table 11b). The median absolute beta change was 0.002 and the median OR/RRR ratio change was approximately 0.001 from the main estimate. Nevertheless, there were 12 changes in FDR significance and 4 direction changes. This suggests that while point estimates were largely robust, some statistical significance conclusions were sensitive to the smaller complete-case covariate sample.

#### 4.1.iii Results of sensitivity analysis removing partial-case inclusion of depression group.

Finally, another sensitivity analysis was undertaken which excluded the partial-case inclusion of depression trajectories (Supplementary Table 11c). This removed 48 participants from the persistent/recurrent trajectory group. This resulted in a median absolute beta change was 0.007 and the median OR/RRR ratio change was 0.029 from the main estimate, indicating that the inclusion of unambiguously classifiable partial cases had minimal influence on the study findings.
